# The acute effects of aerobic exercise on sensorimotor adaptation in chronic stroke

**DOI:** 10.1101/2020.12.10.20231043

**Authors:** Christopher P Mackay, Sandra G Brauer, Suzanne S Kuys, Mia A Schaumberg, Li-Ann Leow

## Abstract

Sensorimotor adaptation, or the capacity to adapt movement to changes in the moving body or environment, is a form of motor learning that is important for functional independence (e.g., regaining stability after slips or trips). Aerobic exercise can acutely improve many forms of motor learning in healthy adults. It is not known, however, whether acute aerobic exercise has similar positive effects on sensorimotor adaptation in stroke survivors as it does in healthy individuals.

**Purpose:** The aim of this study was to determine whether acute aerobic exercise promotes sensorimotor adaptation in people post stroke.

**Methods:** A single-blinded crossover study. Participants attended two separate sessions at the university campus, completing an aerobic exercise intervention in one session and a resting control condition in the other session. Sensorimotor adaptation was assessed before and after each session. Participants were twenty people with chronic stroke. Intervention completed was treadmill exercise at mod-high intensity for 30 minutes.

**Results:** Results demonstrated that acute aerobic exercise in chronic stroke survivors significantly increased sensorimotor adaptation from pre to post treadmill intervention.

**Conclusion:** These results indicate a potential role for aerobic exercise to promote the recovery of sensorimotor function in chronic stroke survivors.

## Introduction

Change in the brain due to neuroplasticity is a foundation principle underpinning the rehabilitation of sensorimotor function following stroke. Plastic effects of acute aerobic exercise on the brain have been previously demonstrated in many domains of cognition [1]. Evidence suggests that the cognitive performance of an individual can be significantly improved due to acute aerobic exercise and that this improvement may be further enhanced when exercising at a moderate to high intensity compared with a lower intensity [2, 3]. Older adults (age 65 years or older) are at an increased risk of cognitive decline, yet there appears to still be benefits of acute aerobic exercise on cognition (executive function), with more demanding cognitive tasks often more sensitive to the effects of physical exercise than less demanding cognitive tasks [4]. As well as improving brain performance from a cognitive function perspective, there is also evidence for specific improvements in sensorimotor function following acute aerobic exercise. This finding corroborates the work of others who have demonstrated that acute exercise improves various forms of motor learning in young healthy adults [5-9], although not all studies show that exercise improves motor learning [10-12]. In a recent study, young healthy participants who first exercised at a moderate intensity on a cycle ergometer for 25 minutes subsequently demonstrated improved movement accuracy and reaction time in a sensorimotor adaptation task [9]. In studies of sensorimotor adaptation, the sensory feedback of a movement is experimentally perturbed, leading to a discrepancy between the predicted sensory outcome and the actual sensory outcome (sensory prediction error) [13]. A perturbation can be created by rotating the visual feedback of a hand movement in a 30° clockwise direction during a target reaching task. Perturbations lead to discrepancies between the predicted sensory feedback about the movement and the actual sensory feedback received (i.e., sensory prediction errors) [14]. With repeated reaches, this sensory prediction error triggers an updating of the motor command to reduce the sensory prediction error, and this learning occurs in an implicit, automatic way. Perturbations also often lead to discrepancies between the predicted task outcomes (e.g., hit the target) and the actual task outcomes (e.g., fail to hit the target), termed task errors [15-18]. Such task errors are thought to promote the use of explicit strategies, to obtain desired task outcomes. Quantifying how quickly individuals return sensory prediction errors and task errors to pre-perturbation levels measures the brain’s ability to learn to adapt to altered sensory feedback via implicit and explicit learning mechanisms, and may indicate the neuroplastic capacity of the individual [19].

Although increasing evidence demonstrates beneficial effects of exercise on motor learning in young, healthy individuals, far fewer studies have tested the acute effects of exercise on motor learning in cohorts with neurological deficits. Isolated studies have demonstrated improvements in motor skill learning in people with Parkinson’s disease following a single session of moderate intensity (60-70% V□O_2_max) cycling [20], and in people with stroke following high intensity interval training [21]. In contrast, one study in stroke patients showed that acute exercise did not improve split-belt treadmill adaptation, which is a form of sensorimotor adaptation involving adaptation to different treadmill speeds for different legs [22]. To the best of our knowledge, the acute effect of moderate to high intensity aerobic exercise on sensorimotor adaptation with goal-directed reaching of the upper limbs in stroke is not known.

Therefore, the aim of this study was to determine if a single bout of moderate to high intensity aerobic exercise performed by people following stroke improved sensorimotor adaptation compared to a control period of rest. We hypothesised that acute aerobic exercise would improve the sensorimotor adaptation of individuals post stroke.

## Methods

### Participants

Twenty people with chronic stroke participated in the study (aged 61 ± 13 years; 76% male). To be included in the study, participants had to be diagnosed with a stroke at least 3 months prior, be able to walk with or without an aid for at least 10 metres and be able to understand three-stage commands. Individuals were excluded if they were unable to walk independently prior to the current stroke, had co-morbidities that might limit their walking (such as arthritis), had an unstable cardiac status, or were unable to understand instructions or provide informed consent.

A sample size of 20 was calculated from data showing changes in BDNF levels after a single bout of exercise in people with MS [23, 24]. Both studies found a change of BDNF levels of 5ng/ml from pre to post, with a maximum SD of 5ng/ml. A sample size of 18 is required to detect a difference of 5+/-5ng/ml at a two sided 0.5 significance level with a power of 80%. The primary outcome measure for the current data set was BDNF, but the results for the analysis of BDNF are currently incomplete and not presented here.

Information collected to describe the sample participants included date, location and type of stroke, age, sex, medical co-morbidities and current medications. Ethical approval for the study was obtained through the local human research ethics committee. All participants provided written informed consent in accordance with the Declaration of Helsinki.

### Experimental design

In a cross-over design, each participant was pseudo-randomised to complete an intervention condition (Treadmill) or control condition (Rest) first, returning at least one week later (washout period) to complete the alternate condition. A minimum washout period of one week was used to allow for any physiological changes due to exercise to return to resting levels, but to minimise any stroke recovery-associated changes to function. Participants were asked to refrain from exercising in the 24 hours prior to their visit. At each visit, participants completed the sensorimotor adaptation task on two occasions, pre and post intervention, resulting in four assessment timepoints (PreControl, PostControl and PreTreadmill, PostTreadmill). The post intervention assessment aimed to commence within 15 minutes of completing the treadmill or rest condition.

### Intervention

The intervention condition consisted of a single session of moderate-high intensity aerobic exercise (65% of heart rate reserve) for 30 minutes walking on a standard treadmill (Landice L7 treadmill), with arms placed comfortably by participants on the rail (in front) or swinging freely. Information describing the specific features of the training session (e.g. heart rate, blood pressure response, treadmill speed, total distance walked) were recorded to monitor the response to exercise and adherence to the exercise protocol. The single treadmill session included a progressive increase in intensity (usually increased speed, or gradient) to reach the target heart rate (approx. 5 mins) as well as a cooling down period (approx. 5 mins) to allow for participants to return towards resting levels for vital observations. The warm-up and cool-down were included as part of the total 30 minutes of walking exercise. Target heart rates were calculated using the Karvonen method [25] with levels adjusted for those taking heart rate lowering medications (i.e. beta blockers), following methods previously published in post stroke populations [26, 27]. Heart rate was measured using a chest strap monitor (polar-electro) and monitored by a research assistant who facilitated changes in treadmill parameters to enable participants to reach their target. Participants were asked to self-rate their intensity of exercise every 10 minutes verbally using BORG’s 6-20 scale rating of perceived exertion [28]. Participants were instructed to walk at a pace that resulted in a rating between 11 (fairly light) and 14 (somewhat hard) on the scale. The control condition involved an equivalent time period (30 minutes) of seated resting where participants were provided with an education session about the impact and effects of stroke by the same research assistant.

### Sensorimotor adaptation task

#### Apparatus and setup

Participants were seated in front of a desk (approximately 50 cm from their coronal plane) and asked to move a digitising pen (15.95 cm long, 1.4 cm wide, 17 g) on a digitizing tablet (WACOM Intuos4 PTK 1240, size: 19.2 x 12 in., resolution = 0.25 mm) from an origin to a target point. The pen’s position on the tablet (XY coordinates) was sampled at 100 Hz and displayed in real time as a circular cursor with a 5-pixel radius (1.25 mm) on a horizontally placed computer monitor. Direct vision of the hand was prevented by placing the tablet and the hand directly beneath an opaque stand, with the horizontal monitor placed atop the stand.

#### Task instructions

Participants first received task instructions to move an on-screen cursor from the start to the target, in a straight line, in a single movement, as quickly and as accurately as possible. Further, participants were instructed that the feedback of the movement would be changed from time-to-time, and that participants were to change their movement in response to this change of feedback, whilst keeping movements as straight as possible.

In each trial, the participants’ task was to move from the origin location through the target location as quickly and accurately as possible using their dominant upper limb. Targets were presented in one of three locations (210, 225 or 240 degrees from the right horizontal plane) in random order. These target directions were selected such that target-reaching movements involved the horizontal adductors of the shoulder joint. After moving through the target, or past the target, a high-pitched tone sounded to indicate trial completion. Following completion of a trial the origin location was re-displayed immediately and participant directed to repeat task. If the participant did not complete a successful trial, they were directed to return to the origin location and repeat the trial.

First, participants encountered 18 baseline trials under normal (correct) feedback with no rotation. Participants then immediately completed 66 adaptation trials, where the visual feedback on the display monitor was perturbed by rotating it 30 degrees in a clockwise direction (1^st^ testing day) or counter-clockwise direction (2^nd^ testing day). After the adaptation block, to notify participants that the perturbation had been removed, a popup dialog box appeared with the statement “In the next few trials, the disturbance that the computer applied would be removed. Please aim straight to the target.” The instructions on-screen were read out by the experimenter to ensure it was understood. This was immediately followed by 6 no-feedback trials. Finally, participants completed 36 washout trials under normal cursor feedback conditions (i.e., no cursor rotation), to return behaviour to an unadapted state.

### Data Processing and Analysis

Custom scripts written in LabVIEW scored reach directions, which were quantified at the 15^th^ data point (150 milliseconds into the reach), as online movement corrections typically occur after 150 ms into a reach. Trials with reach direction outside a 120 degree range of the target (60 degrees on either side of the target) were discarded as outliers [29]. Trials were binned into cycles of one visit to each of the three targets. The dependent variable was percent adaptation [30, 31], which quantified reach directions in every cycle relative to the ideal reach direction by calculating reach directions as a percentage of ideal reach directions resulting from perfect adaptation performance. Ideal reach direction was 30 degree clockwise for a 30 degree counter-clockwise rotation, and 30 degrees counter-clockwise for a 30 degree clockwise rotation.

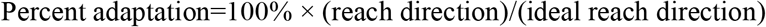

As observed previously [9, 31], there was a rapid error reduction phase of reaching where a majority of learning occurred followed by a slower rate of adaptation. Here, as the targets were spaced close together, rapid error reduction occurred in trials 1-9 (i.e., the first three cycles), similar to recent work [32]. At completion of the fourth cycle, adaptation was greater than 70% both before and after the treadmill and control conditions. We thus selected the first three cycles to quantify rapid error reduction.

Individual differences in reach directions at baseline can affect measures of adaptation [33]. Previous methods of accounting for this by subtracting pre-perturbation behaviour from post-perturbation behaviour is more sensitive to noisy baselines and risk of Type 2 error (Vickers, 2001; Vickers, 2014). To account for pre-perturbation baseline biases, we entered percent adaptation averaged from the final three cycles before rotation onset (i.e., percent adaptation in the last five baseline cycles) as covariates in all of our analysis of covariance analyses.

After the adaptation trials, there were two cycles where no feedback was provided to the participant. That is, unlike all previous trials where participants could follow a tracking cursor with their vision, the participants received no real time feedback about where they were reaching. The first no-feedback cycle was taken as a measure of implicit learning, similar to previous work [18].

Our outcome measures of interest were (1) adaptation performance (2) implicit aftereffects, quantified as reaches that remained adapted despite notification of perturbation removal in the no-feedback block and (3) explicit learning, estimated as the volitional disengagement of adapted behaviour after receiving notification of perturbation removal (i.e., the change in percent adaptation from the mean of the last three adaptation cycles to the first no-feedback cycle after receiving notification that the perturbation was gone), and (4) de-adaptation performance. To evaluate these measures, we ran ANCOVAs with the between-subjects factor Intervention Order (Control First, Treadmill First) and the within-subjects factors: Intervention (Control, Treadmill), Time (Pre-Intervention, Post-Intervention) and (where applicable) Cycles (cycles 1..3), with pre-rotation biases as covariates of no interest (estimated from mean percent adaptation in the last three baseline cycles). Where appropriate, Greenhouse-Geisser corrections were applied. Alpha was set at 0.05. SPSS v24.0 was used for statistical analyses.

## Results

### Reaching with rotated feedback (adaptation trials)

During the rapid error reduction phase of the adaptation trials (cycles 1-3), a significant Intervention x Time interaction was observed [F (1,13) = 6.399, p = 0.027, partial eta squared = 0.346]. Follow-up ANCOVAs were run separately for the Treadmill and Control interventions. Percent adaptation increased pre-to-post in the Treadmill intervention [Figure 2B, significant main effect of Time, F(1,15) = 6.241, p = 0.025, partial eta-squared = 0.247, PreTread 52.5 ± 6.2% [39.1, 66.0] vs. PostTread 65.7 ± 3.8%) [57.4, 74.0], but not for the Control intervention [Figure 2A, non-significant main effect of Time, F(1, 15) = 1.138, p = 0.303, partial eta-squared = 0.07, PreControl 62.8 ± 5.9% [49.9%, 75.9%] vs. PostControl 53.9 ± 6.1% [40.5%, 67.5%]].

**Figure 1.**
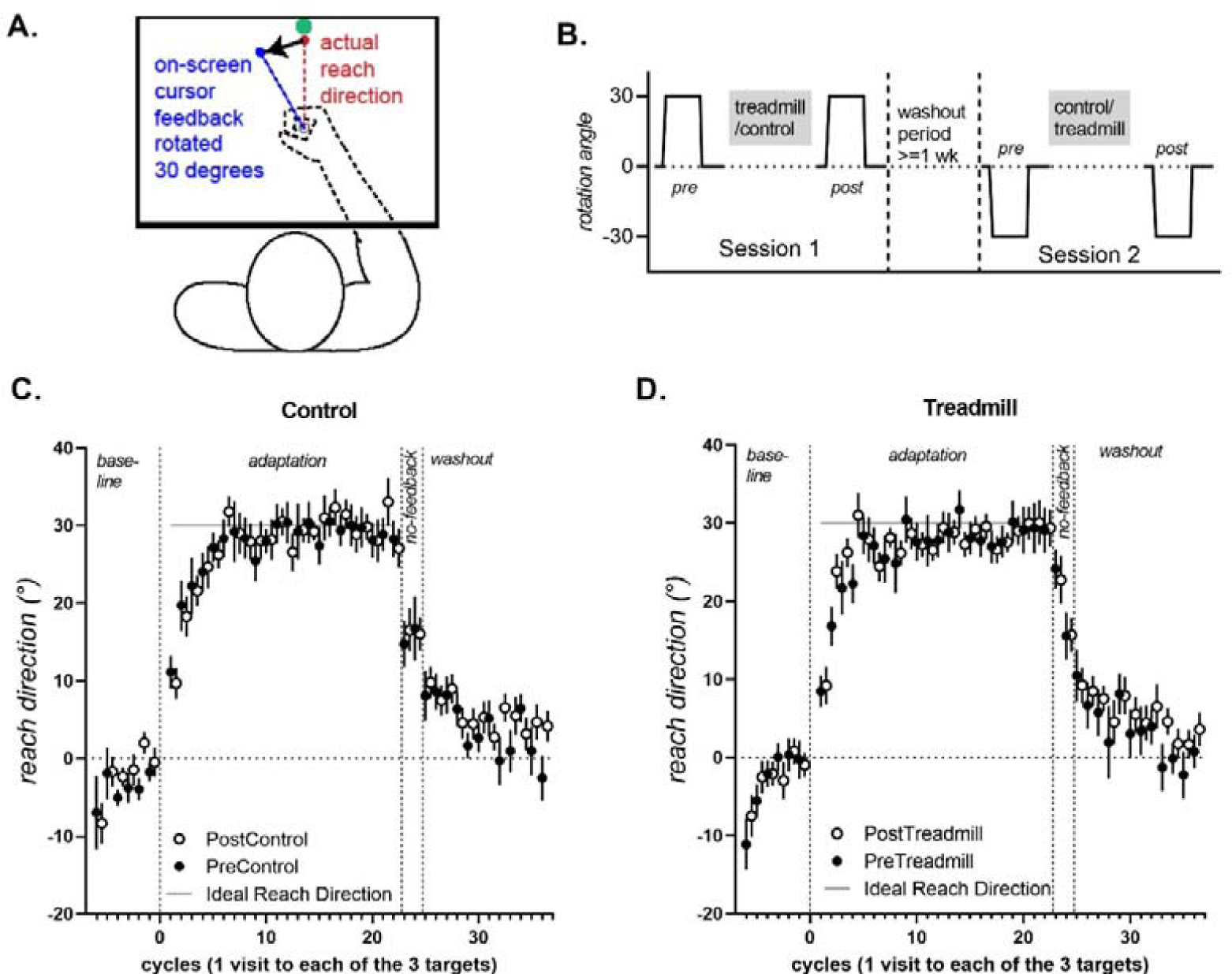
A. Experimental set-up. Vision of the arm is occluded as participants make reaching movements on a digitizing tablet placed under a stand: cursor feedback of the hand is shown on a monitor screen placed horizontally on the stand. B. All participants completed Session 1 with a 30 degree clockwise rotation of cursor feedback, and the ideal reach direction was 30 degrees clockwise. On Session 2, participants encountered a 30 degree counter clockwise rotation, and the ideal reach direction was 30 degrees clockwise. C&D. Reach direction for each cycle (1 cycle= 1 visit to each of the 3 targets), for the control and treadmill conditions. For ease of comparison, datasets which experienced a clockwise rotation (and had a - 30 ideal reach direction) were sign-flipped for this figure, such that reaches that are close to +30 represent better adaptation for all datasets. Data from the Post conditions are offset by 0.5 to avoid being obscured by data from the Pre conditions.

**Figure 2:**
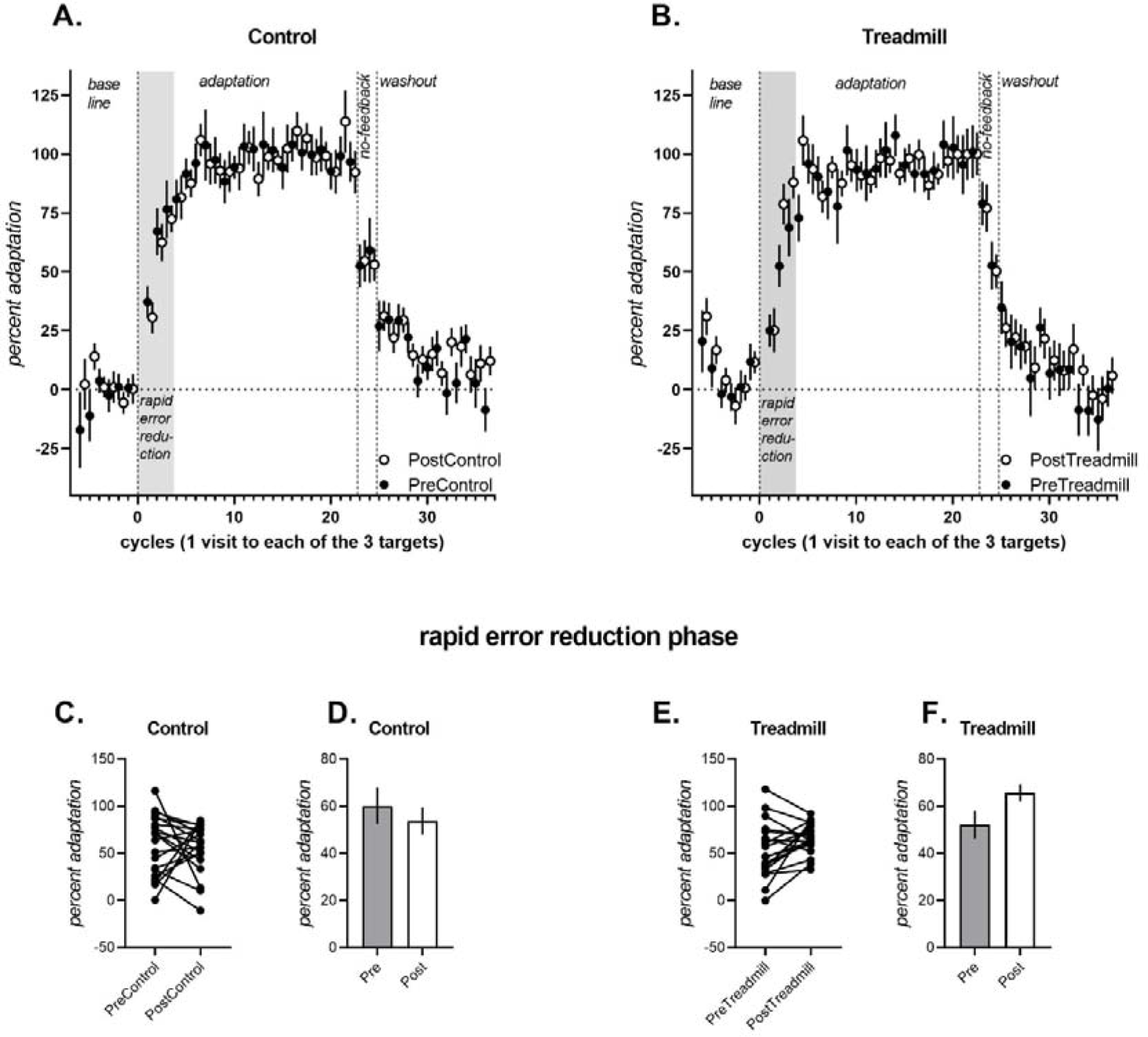
Percent adaptation of participants during the adaptation (cycles 1-22), no-feedback (cycles 23-24) and washout (cycles 25-36) trials for the control condition (A) and the exercise condition (B). Note that data from the Post conditions are offset by 0.5 to avoid being obscured by data from the Pre conditions. There was a significant interaction between the control and exercise condition from pre to post in the rapid error reduction phase (shaded region, i.e., first 3 adaptation cycles). Individual participant data in the rapid error reduction phase is shown for the control condition (C) and the treadmill condition (E). Covariate adjusted mean percent adaptation for the rapid error reduction phase is shown for the control (D) and treadmill (F) conditions

### Implicit aftereffects and Explicit learning

Implicit learning (Figure 3.A), measured as percent adaptation in the first no-feedback cycle after receiving notification of the perturbation removal [similar to 16], was larger overall in the treadmill intervention conditions than the control intervention conditions, but did not differ significantly from pre-to post-between control and treadmill conditions, as the Intervention x Time interaction was not significant, (p > 0.05). The main effect of treadmill and the main effect of Time was also not significant.

**Figure 3.**
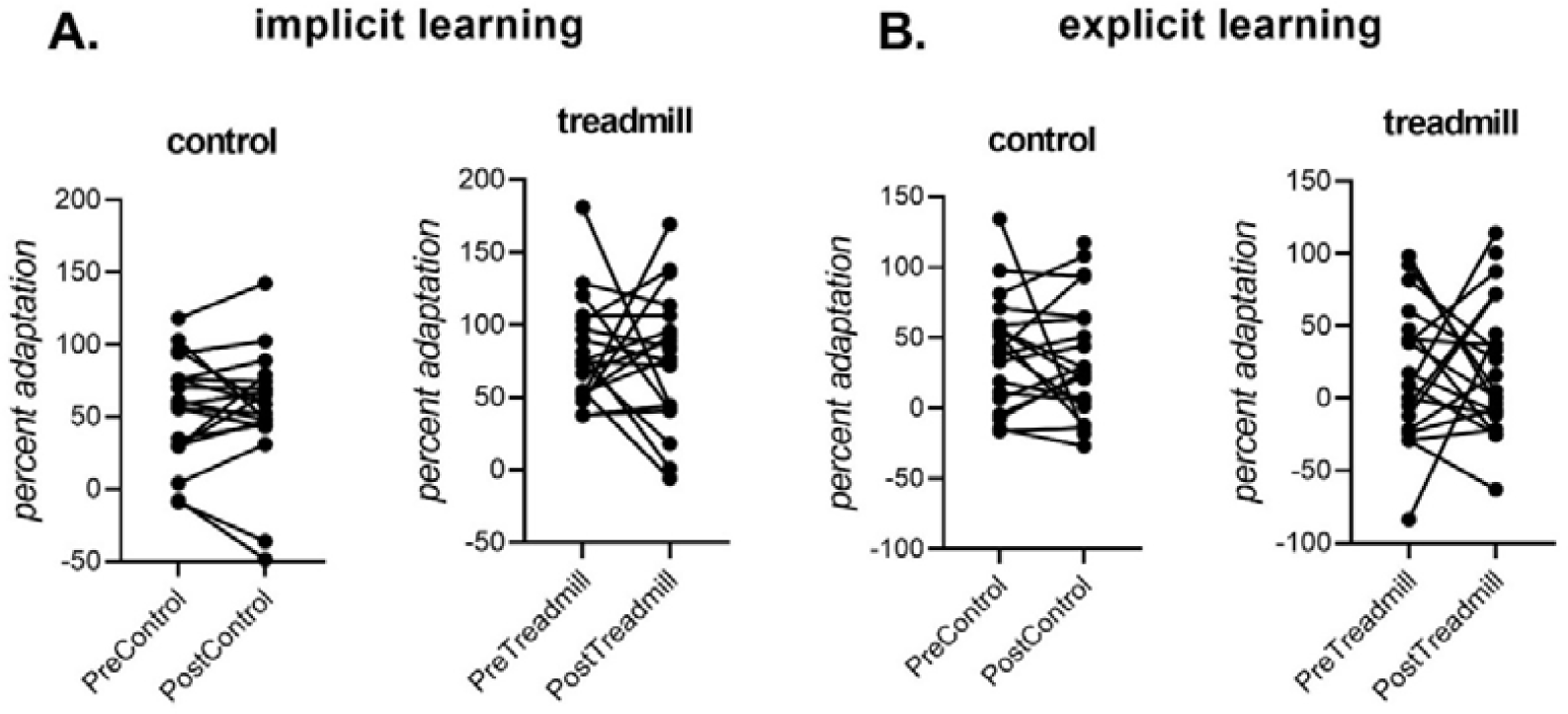
A. Individual participant data for implicit learning, measured as percent adaptation in the first no-feedback cycle (i.e., movements that remained adapted despite being notified that the perturbation had been removed). B. Individual participant data for explicit learning, measured as the change in percent adaptation upon notification of the perturbation removal). Implicit and explicit learning did not differ significantly from pre-to post-control and/or pre-to-post intervention.

All conditions showed a clear change in reach directions after receiving instructions that the perturbation had been removed (see Figure 2), indicating a role for explicit learning in this sensorimotor adaptation task. Explicit learning did not differ reliably between before and after the control and treadmill interventions (all main effects of Intervention and Time, and all interactions with Intervention and Time p > 0.05).

### Reaching with normal feedback (de-adaptation trials)

Pre and post-intervention de-adaptation did not differ reliably between Control and Treadmill intervention conditions, [non-significant main effect of Intervention, non-significant x Time interaction, F(1,12) = 0.4, p = 0.538, partial η-squared = 0.03). Participants in both intervention conditions returned to a normal level of baseline reaching, as expected.

## Discussion

This study found that a single bout of moderate to high intensity aerobic exercise was sufficient to increase sensorimotor adaptation performance of chronic stroke survivors. The improvement in reaching performance (observed as a faster rate of adaptation post-intervention compared to pre-intervention) was observed during the rapid error reduction phase of the trials after the treadmill exercise had been performed, but not after the rest condition. Thus, the present study showed that a single session of moderate to high intensity aerobic exercise increased capacity to improve sensorimotor adaptation in chronic stroke survivors.

Immediate improvement in reaching performance post-exercise was specifically seen in the rapid error reduction phase of adaptation. Participants demonstrated increased ability to adapt to the imposed rotation in the first nine trials following the exercise compared to the control condition. Such an improvement in reaching performance following exercise may represent an acute change in the brain leading to an enhanced internal environment that could facilitate adaptation. We note that the current findings contrast with some recent work by Charalambous et al. (2018, 2019) who found no effects of exercise on consolidation in locomotor adaptation in stroke patients [12] and in healthy controls [10]. Several differences in the design might have led to this pattern of results. First, although locomotor adaptation is also susceptible to effects of explicit learning, people do not appear to actively engage explicit learning in locomotor adaptation [34]. In contrast, behaviour in the type of reach adaptation paradigm used here has a large contribution from explicit learning, particularly in the early phases of learning [35]. Second, Charalambous et al. (2018, 2019) examined the effects of exercise on consolidation of adaptation by testing retention after a >= 24 hour delay, in contrast to the current work which retested participants immediately after the exercise/control interventions. Finally, the duration of exercise (≈5 mins) in Charalambous et al. (2018, 2019) was shorter than that used here (≈25 mins). Future studies explicitly measuring the effects of these factors will be important for designing exercise-based interventions in movement rehabilitation.

The current finding that exercise has immediate effects on improving early adaptation in stroke patients is consistent with recent work in young healthy adults. Neva et al. (2019) demonstrated that in young, healthy individuals, performance of a visuomotor upper limb rotation task improved (reflected by a lower peak lateral displacement) following a single bout of aerobic cycling exercise (performed at a similar intensity and duration to the current study, 65-70% of max HR for 25 minutes). This improvement occurred immediately after exercise and was retained at 24 hours post-exercise. Acute benefits of moderate to high intensity exercise have also been shown following treadmill running and high intensity shuttle running with improvements to a motor learning task and a visuomotor adaptation task respectively in healthy young adults [36, 37]. These studies collectively strengthen the argument that completing exercise prior to a movement task (sensorimotor or visuomotor) has a positive impact on performance. A number of mechanisms might contribute to exercise-related improvements in learning. This improvement may be related to changes in the brain that accompany intense exercise, such as upregulation of neurotrophic factors such as brain derived neurotrophic factor (BDNF), or proteins such as growth hormone, increased cortisol levels and/or changes to neurotransmitter release [38-40]. Other potential mediators for improved performance include the salience of the task at hand (i.e. how relevant/important is the task to the individual completing it) and the ability of the client to engage with and concentrate on the task. These two mediators (salience and concentration) are of particular importance, highlighted by stroke guideline statements that encourage active task practice outside of scheduled therapy hours to maximise functional return in rehabilitation [41].

Evidence supports the prescription of moderate to high intensity aerobic exercise to improve sensorimotor performance in healthy individuals. The present study extends these findings to demonstrate that in stroke survivors, utilising moderate to high intensity exercise can improve sensorimotor adaptation. Having a positive change in performance (measured as an increased adaptation) may represent an exercise induced change in the brain. It has been previously stated that BDNF concentrations increase following a single bout of aerobic exercise [39]. BDNF is a neurotrophin that plays a key role in the formation of new neurons, development and strengthening of existing neurons and in the restructure of the neuron pool with use [42-44]. It may be possible to harness increased levels of BDNF following exercise to enhance sensorimotor performance, and we have shown that increased levels of BDNF can occur in response to a program of aerobic exercise in neurological populations [45]. This is an important concept for chronic stroke survivors due to the stagnant nature and plateau that often accompanies this phase of recovery. Providing stroke survivors with a way to augment their recovery could lead to enhanced motor re-adaptation, functional gains and ultimately boost community participation of this group.

Average time post stroke for participants in this study was four years, representing the chronic phase of recovery. We note that although gains are still made in this phase of recovery, the magnitude of gains may be less than in a more acute recovery phase [46]. The majority of post-stroke recovery is generally observed to occur within the acute (1 - 7 days) and subacute phases of recovery (7 days – 6 months) [47]. Harnessing potential changes in the brain due to exercise is important for all stages of recovery post stroke, but the magnitude and impact of this change may be increased in the earlier phases of recovery. It is therefore important to consider the timing of an aerobic exercise intervention post-stroke to maximise patient benefit in this way. Implementing an aerobic exercise intervention during the subacute phase of recovery may represent a more appropriate temporal window for enhancing neuroplastic benefit. Stroke survivors in the chronic phase of recovery, however should still be encouraged to participate in aerobic exercise for potential benefit.

As this study was a within-subject (crossover) design, participants acted as their own control. Participants engaged in the sensorimotor adaptation task four times, on two separate testing days, with a one-week washout period between the two testing days. Compared to the control intervention, the treadmill exercise intervention increased pre-to-post intervention improvement in the rapid error reduction phase. There is the possibility that carryover effects between testing days might have partly contributed to the current pattern of results. We did, however, attempt to control for this with an equal and opposite rotation on the second testing day in a counter-balanced order (30 degrees clockwise on Day 1 vs 30 degrees counterclockwise on Day 2). We also randomised the number of participants who completed the control condition on their first visit compared to the treadmill condition. Additionally, assessors were blinded to the intervention completed at each session to eliminate bias. There was no follow up testing of the participants to examine whether they retained an improvement due to exercise at a later date. Therefore, we cannot comment on the impact that the exercise may have had on longer-term learning. The assessment item to quantify motor learning in this study was a sensorimotor adaptation task. Although this task has been widely used, it might not fully describe real-life motor skill acquisition [48]. This sensorimotor adaptation task, however, enabled the sensitive detection of exercise-related improvements in motor learning here.

This is the first study to our knowledge to demonstrate an improvement in sensorimotor adaptation following moderate-high intensity exercise in a stroke population. Due to the neurological deficit incurred by stroke survivors, being able to improve adaptation of a motor task through aerobic exercise has the potential to significantly improve function in this cohort.

**Table 1:** Exercise performance data (n = 20)

|  | Mean & St. Dev |
| --- | --- |
| **<u>TREADMILL DATA</u>** |  |
| **Speed Maximum (km/hr)** | 3.94 ± 1.54 |
| **Time (mins)** | 28.58 ± 3.46 |
| **Distance (metres)** | 1761.2 ± 660.8 |
| **Maximum predicted HR (bpm)** | 157 ± 16 |
| **HR Max achieved (bpm)** | 122 ± 19 |
| **% of Max HR** | 78.3 ± 13.6 |
| **Modified Borg Maximum** | 13 ± 1 |

Legend: HR - Heart Rate, bpm - beats per minute

**Table 2:** Descriptive results (n = 20)

|  |  |
| --- | --- |
| **Average age** | 60 years ± 14 |
| **Time since stroke** | 3 years, 11 months |
| **Male participants** | 76% |
| **10 Metre Walk Time** | 8.5 seconds |
| **Dominant hand used for reach** | 17/20 or 85% |

## Data Availability

All data obtained in the study is available on request.

## Acknowledgements

No funding was provided for this study. The authors would like to acknowledge the in kind contributions received from the School of Human Movement and Nutrition Science at The University of Queensland, and the School of Health and Rehabilitation Sciences at The University of Queensland in the form of utilisation of resources, buildings and assistance from staff, including Katrina Kemp.

